# Robustness of a Restriction Spectrum Imaging (RSI) quantitative MRI biomarker for prostate cancer: assessing for systematic bias due to age, race, ethnicity, prostate volume, medication use, or imaging acquisition parameters

**DOI:** 10.1101/2024.09.10.24313042

**Authors:** Deondre D Do, Mariluz Rojo Domingo, Christopher C Conlin, Ian Matthews, Karoline Kallis, Madison T Baxter, Courtney Ollison, Yuze Song, George Xu, Allison Y Zhong, Aditya Bagrodia, Tristan Barrett, Matthew Cooperberg, Felix Feng, Michael E Hahn, Mukesh Harisinghani, Gary Hollenberg, Juan Javier-Desloges, Sophia C. Kamran, Christopher J Kane, Dimitri Kessler, Joshua Kuperman, Kang-Lung Lee, Jonathan Levine, Michael A Liss, Daniel JA Margolis, Paul M Murphy, Nabih Nakrour, Michael A. Ohliger, Thomas Osinski, Anthony James Pamatmat, Isabella R Pompa, Rebecca Rakow-Penner, Jacob L Roberts, Karan Santhosh, Ahmed S Shabaik, David Song, Clare M Tempany, Shaun Trecarten, Natasha Wehrli, Eric P Weinberg, Sean Woolen, Anders M Dale, Tyler M Seibert

## Abstract

**Introduction:** Prostate multiparametric magnetic resonance imaging (mpMRI) has greatly improved the detection of clinically significant prostate cancer (csPCa). However, the limited number of expert sub-specialist radiologists capable of interpreting conventional prostate mpMRI is a bottleneck for universal access to this healthcare advance. A reliable and reproducible quantitative imaging biomarker could facilitate implementation of accurate prostate MRI at clinical sites with limited experience, thus ensuring more equitable patient care. Restriction Spectrum Imaging restriction score (RSIrs) is an MRI biomarker that has shown the ability to enhance the qualitative and quantitative interpretation of prostate MRI. However, patient-level factors (age, race, ethnicity, prostate volume, and 5-alpha-reductase inhibitor (5-ARI) use) and acquisition-level factors (scanner manufacturer/model and protocol parameters) can affect prostate mpMRI, and their impact on quantitative RSIrs is unknown.

**Methods:** RSI data from patients with known or suspected csPCa were collected from seven centers. We estimated effects of patient and acquisition factors on prostate voxels overall (Method 1: benign patients only) and on only the maximum RSIrs within each prostate (RSIrs_max_; Method 2: benign and csPCa patients) using linear models. We then tested whether adjusting for any estimated systematic biases would improve performance of RSIrs for patient-level detection of csPCa, as measured by area under the ROC curve (AUC).

**Results:** Using both Method 1 and Method 2, we observed statistically significant effects on RSIrs of age and acquisition group (*p < 0.05*). Prostate volume had significant effects using only Method 2. All of these effects were small, and adjusting for them did not improve csPCa detection performance *(p ≥ 0.05*). AUC of RSIrs_max_ for patient-level csPCa detection was 0.77 (95% CI: 0.75, 0.79) unadjusted, compared to 0.77 (0.76, 0.79) and 0.74 (0.72, 0.76) after adjustment using Method 1 and 2 respectively.

**Conclusion:** Age, prostate volume, and imaging acquisition factors may lead to systematic differences in RSIrs, but these effects are small and have minimal impact on performance of RSIrs for detection of csPCa. RSIrs can be used as a reliable biomarker across a wide range of patients, centers, scanners, and acquisition factors.

## Introduction

The use of multiparametric magnetic resonance imaging (mpMRI) in diagnosing and planning treatment for prostate cancer (PCa) has greatly improved the detection and management of clinically significant PCa (csPCa: grade group [GG] ≥2), reducing unnecessary biopsies, overdiagnosis, and overtreatment in men suspected of having csPCa^1–7^. However, limited access to specialized imaging centers and radiologists skilled in interpreting mpMRI has hindered the widespread adoption of this technology^8^. Additionally, racial and socioeconomic disparities in the utilization of mpMRI have prevented certain at-risk populations from benefiting from this technology, and these disparities may be in part attributable to lack of access to expert centers^9–13^. A quantitative biomarker could address these challenges reducing the need for expert subspecialist radiologists, provided the biomarker could be reliable across common patient-level and center-level scenarios.

Unfortunately, conventional mpMRI currently lacks a biomarker that can accurately distinguish between non-csPCa and csPCa without subjective expert radiologist interpretation. The apparent diffusion coefficient (ADC), a quantitative metric derived from diffusion-weighted imaging (DWI) during mpMRI, has shown poor reliability as a quantitative biomarker and is only clinically useful after a suspicious lesion is identified; by itself, it is not accurate for patient-level detection of csPCa^14–16^. ADC’s model is too simplistic, providing only an ensemble average of diffusion properties within the multiple, complex microstructural environments of prostate tissue, limiting the specificity of ADC measurements ^17^. For example, ADC values within tumors exhibit significant overlap with non-malignant conditions in the prostate, like prostatitis and benign prostate hyperplasia (BPH)^18,19^. Variations in pulse sequences and *b*-values specific to individual clinical sites further impede the establishment of a standardized classification threshold for ADC to detect csPCa^20^. Despite these limitations, ADC is the current quantitative DWI metric used in the Prostate Imaging Reporting & Data System (PI-RADS) for detecting csPCa^21^.

Restriction Spectrum Imaging (RSI) is a more sophisticated diffusion MRI approach that improves the qualitative^22^ and quantitative^14,23^ interpretation of prostate MRI. Prostate RSI utilizes a multi-compartment model to characterize four different types of diffusion in living tissues: restricted intracellular, hindered extracellular, free diffusion, and vascular flow^14,24^. A quantitative biomarker called the RSI restriction score (RSIrs) has been validated as an accurate classifier of csPCa at both the voxel and patient levels, outperforming conventional ADC and performing comparably to expert PI-RADS interpretation^14,23^. RSIrs maps accurately pinpoint csPCa and make it more noticeable to non-experts compared to mpMRI alone, facilitating improved precision of targeted cancer treatment^22,24^ (Figure 1). RSIrs provides objective estimates of the probability of csPCa without requiring expert radiologists^16^ and has the potential to enhance the accuracy of csPCa detection in the early stages of PCa treatment planning, in conjunction with current clinical tools. However, a critical step for clinical implementation of any biomarker is understanding whether common patient or image acquisition factors may systematically bias RSIrs.

**Figure 1.**
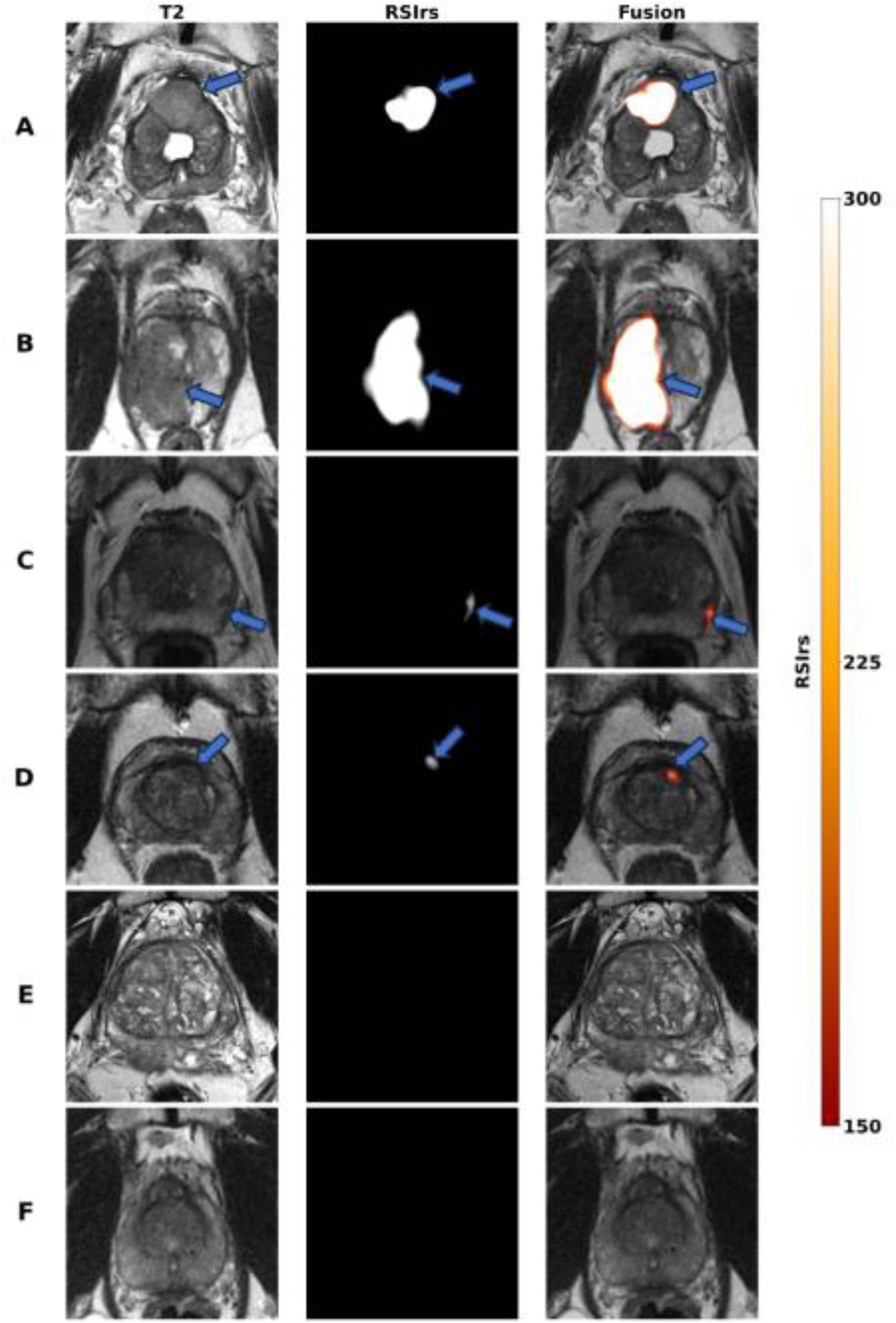
Axial T2-Weighted (T2W) images, RSIrs maps, and RSIrs overlaid on T2W imaging for six patients, illustrating consistent imaging for detection of csPCa despite differences in patient and acquisition factors. Arrows indicate the prostate at the slice location of clinically determined prostate lesions, except for Patients E and F, who had no clinically significant lesions (PIRADS >3). All patients with clinically significant prostate cancer (csPCa) (GG>2) were confirmed by targeted biopsy. The patients are categorized into three groups based on grade group: **High-Risk Group (Patients A and B):** These patients are biopsy-proven GG5, with a PIRADS score of 5, from two different institutions: UCSD and URMC respectively. **Intermediate Group (Patients C and D):** These patients are biopsy-proven GG3, with a large age difference (over 15 years). RSIrs_max_ was between 300 and 350 for both these patients. Patient C had a PIRADS score of 3, and Patient D had a PIRADS score of 5. **Non-csPCa Group (Patients E and F):** These patients have a large discrepancy in prostate size (22 and 129 cubic centimeters, respectively), both with a PIRADS score of 1. RSIrs_max_ was <200 for both these patients. Patient A was biopsy-proven benign, and Patient B was biopsy-proven GG1. The UCSD scan was obtained with a GE scanner, and the URMC scan was obtained with a Siemens scanner. RSIrs_max_ was >500 for both these patients. These images illustrate the similarities in RSIrs_max_ regardless of patient and acquisition factors.

Patient-level factors like age, prostate volume, and the use of 5-alpha-reductase inhibitors (5-ARIs) for treatment of benign prostate hyperplasia (e.g., finasteride and dutasteride) can impact the interpretation of mpMRI^26,27^. Racial and/or ethnic disparities are well documented for PCa, particularly among Black or African American men, although mpMRI appears equally useful for patients of different races and ethnicities with expert interpretation^28,29^. We previously found that changing the echo time (TE) during RSI acquisition has a modest effect on RSIrs that can be effectively accounted for through simple calibration^30^. However, differences in RSIrs that might result from variation in scanner model/manufacturer and other acquisition parameters are not well understood. Our objective in the present study was to assess any significant effect on RSIrs from patient and acquisition factors. Additionally, we aimed to determine whether adjusting for any such effects could improve the detection between csPCa and non-csPCa (i.e., benign prostate or GG 1 PCa).

## Methods

### Population Demographics

Prostate MRI data from 2845 patients with RSI were collected from seven imaging centers belonging to the Quantitative Prostate Imaging Consortium (QPIC). These institutions include the Center for Translational Imaging and Precision Medicine at the University of California San Diego (UCSD CTIPM), Massachusetts General Hospital (MGH) affiliated with Harvard University, UC San Diego Health (UCSD Health), University of California San Francisco (UCSF), University of Rochester Medical Center (URMC), University of Texas Health Sciences Center San Antonio (UTHSCSA), and University of Cambridge. All participating institutions received approval from their respective institutional review boards (IRBs). Prospectively gathered data from UTHSCSA and Cambridge were collected with written consent from participants, while the remaining institutions obtained a consent waiver from their IRBs for retrospective use of clinical records. Clinical records were reviewed to extract PI-RADS scores, biopsy results, and patient-level factors of interest, including age, race, ethnicity, prostate volume, and 5-ARI use.

Male patients over 18 years old who underwent prostate mpMRI with RSI were eligible for inclusion in this study. Exclusion criteria included prior treatment for PCa, metal implants in the pelvis, and lack of available biopsy results within 6 months of a positive mpMRI (PI-RADS ≥3). Non-csPCa patients were defined as those (1) with confirmed benign findings or GG 1 PCa based on biopsy histopathology, or (2) with a non-suspicious mpMRI (PI-RADS 1 or 2) and a prostate-specific antigen density (PSAD) < 0.15. Patients were categorized into acquisition groups based on scanner model/manufacturer and RSI protocol; this generally correlated with imaging center, though some centers replaced scanners or otherwise significantly changed their acquisition protocols over the course of the study and therefore contributed data with more than one acquisition group (Supplementary Table 1). Acquisition groups were formed based on the following criteria: same scanner model/manufacturer, equivalent *b*-values, similar voxel size (within 25%), similar TE (within 10 ms). Any groups with less than 15 patients were excluded. All scans were obtained using 3-tesla MRI scanners (GE Discovery MR750/SIGNA Premier or Siemens MAGNETOM Skyra/Trio) from two manufacturers (GE Healthcare, Waukesha, WI, USA; SIEMENS Healthineers, Erlangen, Germany). Clinical MRI examinations were performed and interpreted following the guidelines of PI-RADS v2/v2.1. Prostate segmentation was conducted using an FDA-cleared artificial intelligence (AI) tool (OnQ™ Prostate -Cortechs.ai; San Diego, CA) which has been previously validated to yield results comparable to manual segmentation by an expert radiation oncologist^31^.

### RSI Data Acquisition and Processing

All post-processing was done using MATLAB (MathWorks; Natick, MA) with custom programs. Post-processing included correction of distortions induced by *B_0_* inhomogeneity, eddy currents, and gradient nonlinearity^17,32^. Noise correction was performed to eliminate bias in the DWI signal stemming from the presence of the noise floor. Linear fitting of the RSI model (Equation 1) to the post-processed DWI data was performed to estimate signal contributions from each of the four RSI model compartments (C_1_: restricted intracellular, C_2_: hindered extracellular, C_3_: free diffusion and C_4_: vascular flow). The RSI-C_1_ compartment signal was normalized by the median DWI signal at b=0 (s/mm^2^) within the prostate to calculate each patient’s voxel-wise RSIrs map.

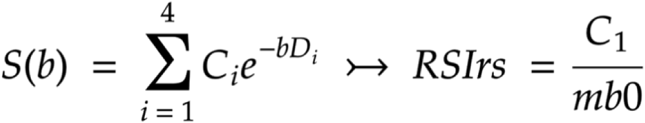

Equation 1: Formula for computing RSIrs where S(*b*) represents the RSI signal based on the *b*-value from diffusion weighted imaging (arbitrary signal units). C_i_ denotes the RSI compartment signal contribution, and D_i_ is the fixed compartmental diffusion coefficient as described by Conlin et al.^24^ mb0 is the median DWI signal at b=0 (s/mm^2^) within the prostate

### Statistical Model Parameters

We estimated systematic effects of the following factors: age, race, ethnicity, prostate volume, 5-ARI use (defined as currently taking this medication or medication ended within 6 months before MRI), and acquisition group (representing MRI scanner manufacturer/model and RSI acquisition parameters). For categorical variables, the reference values were as follows: White Non-Hispanic for race/ethnicity group, no current/recent 5-ARI use, and a common acquisition group at the two UCSD centers (denoted UCSD CTIPM_Discovery1_/UCSDH_Discovery_). We included cancer GG in the model due to the strong association of RSIrs with GG, which is the primary rationale for its utility as a csPCa biomarker. GG was included as a categorical variable with benign as the reference and GG 1, 2, 3, or 4-5 as other values^14,16,33,34^. The maximum RSIrs (RSIrs_max_) in the prostate is a patient-level detector of csPCa and is the most studied application of RSIrs^14,16^. Therefore, we used two methods of modeling effects on RSIrs_max_

### Method 1 (All prostate voxels; only benign and GG 1 cases used to estimate effects)

We used a linear mixed effects model to assess the impact of patient and acquisition factors using voxel-level data from all prostate voxels in a subset of patients were not diagnosed with csPCa. Only patients without csPCa were included to avoid cancer-related RSI effects. Absence of csPCa was determined by either a negative biopsy (benign results or GG1 only) or non-suspicious mpMRI (PI-RADS 1 or 2). To minimize the possibility of occult csPCa affecting model estimation, we additionally excluded any patients without known csPCa if prostate-specific antigen density (PSAD) was ≥0.15. As each prostate contributes many voxels, we included patient case as a random effect in the mixed effects model to account for repeated measures. Method 1 models were fit using fitlme() in MATLAB^35^.

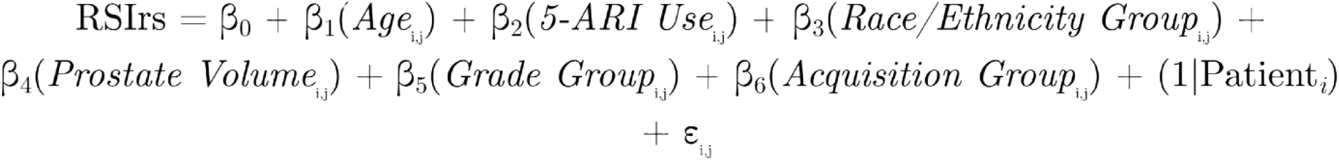

Equation 2. Linear mixed effects model formula to predict RSIrs at the voxel level based on patient and acquisition factors for all prostate voxels. β_0_, β_1_, …, β_5_, β_6_ denotes the respective predictor coefficient estimates, *i* represents the *i-*th patient, *j* represents the *j-*th voxel for the *i*-th patient, (1|Patient) represents the random effect term and ε represents the vector of residual error terms.

### Method 2 (Maximum prostate voxel; all cases used to estimate effects)

We also estimated effects of patient and acquisition factors on only the RSIrs_max_ within each patient’s prostate. Since there was only one RSIrs_max_ value per patient, we employed a multiple linear regression model using fitlm() in MATLAB^36^.

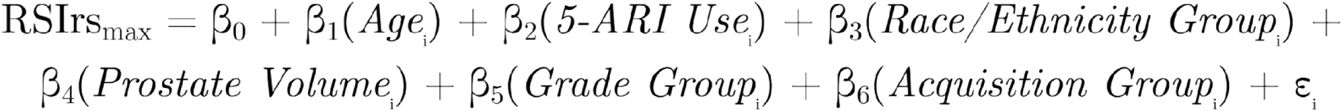

Equation 3. Multiple linear regression model formula to predict RSIrs_max_ within the prostate for a given patient based on patient and acquisition factors. β_0_, β_1_, …, β_5_, β_6_ denotes the respective predictor coefficient estimates, *i* represents the *i-*th patient, and ε represents the vector of residual error terms.

### Impacts on csPCa Detection Performance

To evaluate whether patient and acquisition factors impact the performance of RSIrs_max_ for detection of csPCa, we adjusted RSIrs_max_ for each patient by applying the linear shifts estimated from significant acquisition and patient effects with both methods. We then computed the area under the receiver operating characteristic (ROC) curve (AUC) before and after adjustment to assess the impact of these effects. To account for the uneven distribution of patient and acquisition factors in our dataset, we randomly sampled from a subgroup consisting of only patients with the factors of interest from both methods and matched them with patients without those factors but whose cancer was of the same Grade Group (GG). We stratified our random sampling based on GG to mitigate effects on AUC due to varying proportions of csPCa. Adjustments were made using a linear transformation for each significant factor, as identified by the two different estimation methods. Median differences in AUC and 95% confidence intervals (based on 10,000 bootstrap samples) were compared before and after adjustment for patient and acquisition factor effects. Secondarily, we also re-ran the matching and bootstrapping within subgroups selected only for the statistically significant factors using Method 1 or Method 2, respectively, and compared pre-and post-adjustment within these distinct subgroups.

### Sample Estimation of Significant Effects

It is unknown how many patients are needed to estimate systematic effects on RSIrs_max_ due to patient or acquisition factors. This could influence interpretation of the present study’s statistical power, and it would be helpful to know how many patients might be needed to estimate additional possible effects in the future. To determine the minimum number of patients needed to estimate a patient or acquisition effect, we analyzed patient subgroups with a given factor and compared them to patients from the reference population. For each single factor group, we iteratively bootstrapped 10,000 samples ranging for each of a range of sample sizes, from one to the maximum group size. And for each of these bootstrap sample size groups, we calculated the median coefficient for the factor of interest and the corresponding 95% CI. Then, we determined the mean upper and lower bounds of all 95% CIs across all bootstrap sets and plotted these against sample size to visualize the impact of sample size on the accuracy and precision of the estimated effect for that factor. The minimum sample size to yield a bootstrap median effect estimate within 1% of the coefficient using all data was considered a reasonable threshold for the minimum sample size to estimate acquisition group effects.

## Results

1890 patients met the inclusion criteria (Figure 2). Among them, 94 patients self-identified as White and Hispanic, 1226 as White and Non-Hispanic, 65 as White and Other/Unknown ethnicity, 120 as Asian, 117 as Black, 6 as American Native, and 6 as Native Hawaiian or Other Pacific Islander; race was not reported by 256 patients. 84 patients were currently on 5-ARIs at the time of MRI or had used the medication in the 6 months before their prostate MRI. The median age was 70 with an interquartile range (IQR) of 64-75 years. The median prostate volume was 51 with an IQR of 36-74 mL. One outlier was excluded due to artifact that yielded RSIrs_max_ greater than 15 standard deviations from the population mean (Table 1).

**Figure 2.**
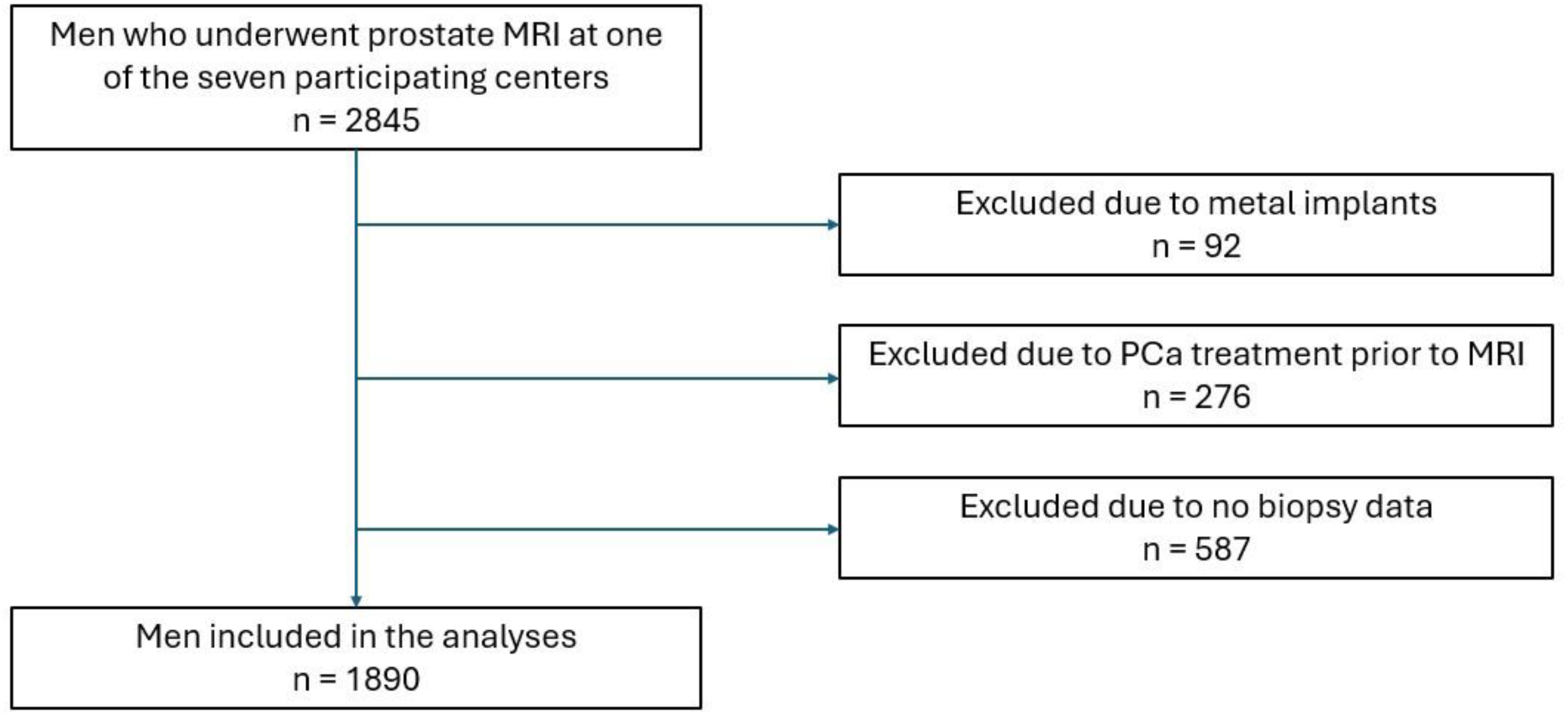
Patient flowchart depicting the selection process of 2845 men over 18 years old undergoing prostate MRI. After applying exclusion criteria, 1890 men were included in the final analysis. 1 additional patient from this dataset was excluded from analysis due to artifact that yielded RSIrs_max_ greater than 15 standard deviations from the population mean.

**Table 1.**
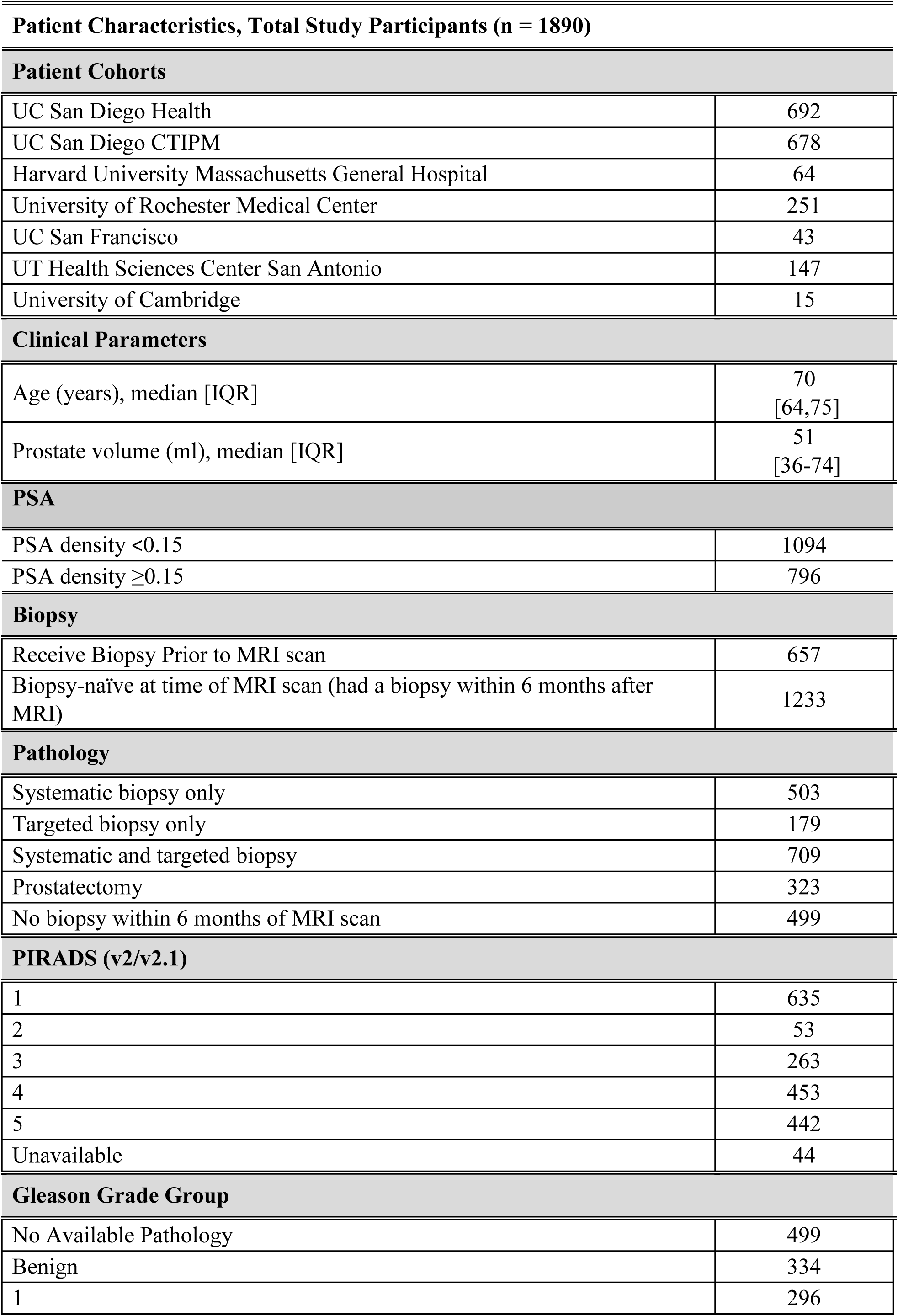
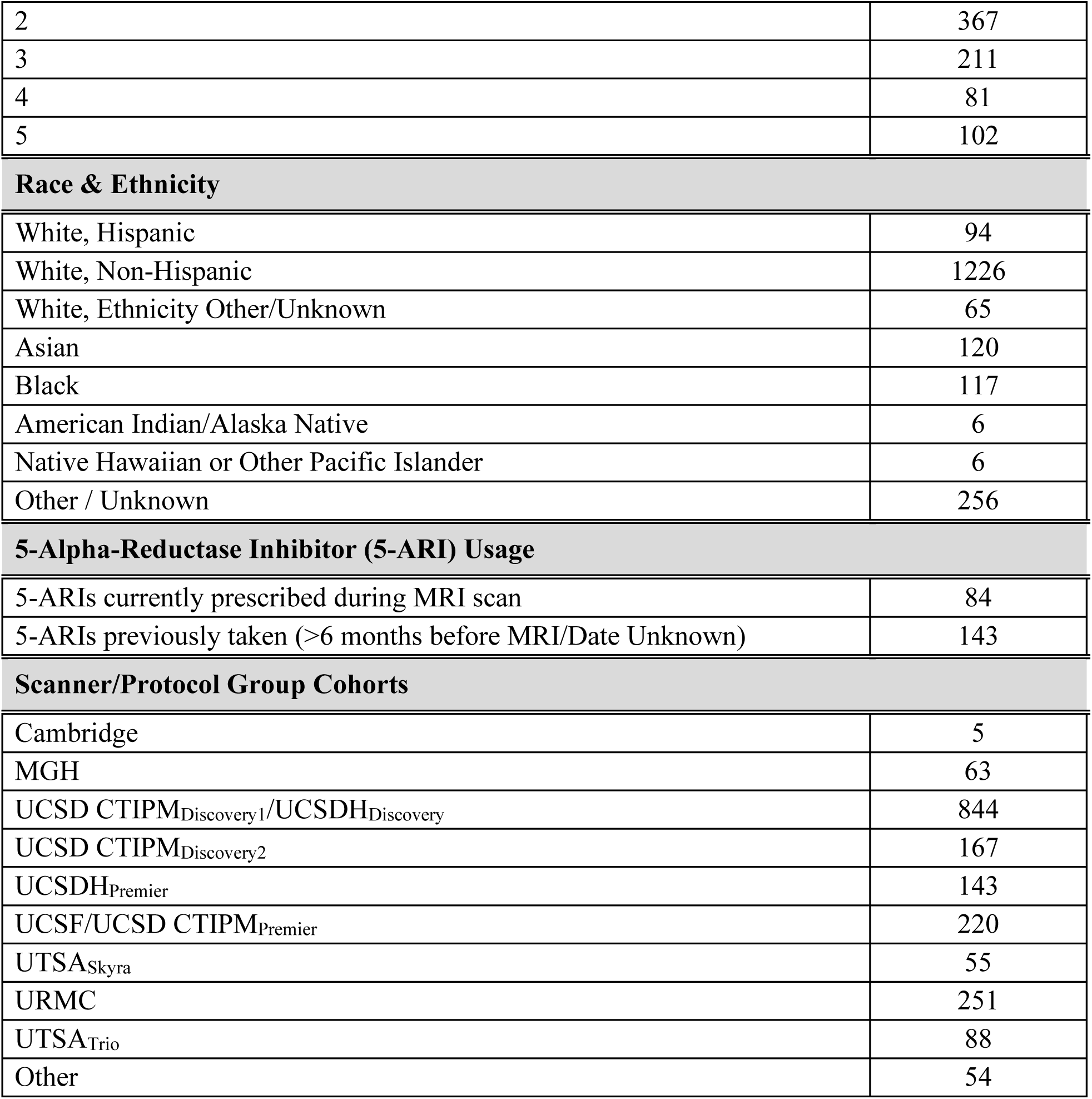
Patient Characteristics. Abbreviations: PSA (Prostate Specific Antigen), Cambridge (University of Cambridge), MGH (Harvard University’s Massachusetts General Hospital), UCSD[H] (University of California San Diego [Health]), CTIPM (Center for Translational Imaging and Precision Medicine), UCSF (University of California San Francisco), URMC (University of Rochester Medical Center), UTHSCSA (University of Texas Health Sciences Center San Antonio)

|  |  |
| --- | --- |
| UC San Diego Health | 692 |
| UC San Diego CTIPM | 678 |
| Harvard University Massachusetts General Hospital | 64 |
| University of Rochester Medical Center | 251 |
| UC San Francisco | 43 |
| UT Health Sciences Center San Antonio | 147 |
| University of Cambridge | 15 |
| **Clinical Parameters** |  |
| Age (years), median [IQR] | 70<br>[64,75] |
| Prostate volume (ml), median [IQR] | 51<br>[36-74] |
| **PSA** |  |
| PSA density <0.15 | 1094 |
| PSA density ≥0.15 | 796 |
| **Biopsy** |  |
| Receive Biopsy Prior to MRI scan | 657 |
| Biopsy-naïve at time of MRI scan (had a biopsy within 6 months after MRI) | 1233 |
| **Pathology** |  |
| Systematic biopsy only | 503 |
| Targeted biopsy only | 179 |
| Systematic and targeted biopsy | 709 |
| Prostatectomy | 323 |
| No biopsy within 6 months of MRI scan | 499 |
| **PIRADS (v2/v2.1)** |  |
| 1 | 635 |
| 2 | 53 |
| 3 | 263 |
| 4 | 453 |
| 5 | 442 |
| Unavailable | 44 |
| **Gleason Grade Group** |  |
| No Available Pathology | 499 |
| Benign | 334 |
| 1 | 296 |
| 2 | 367 |
| 3 | 211 |
| 4 | 81 |
| 5 | 102 |
| **Race & Ethnicity** |  |
| White, Hispanic | 94 |
| White, Non-Hispanic | 1226 |
| White, Ethnicity Other/Unknown | 65 |
| Asian | 120 |
| Black | 117 |
| American Indian/Alaska Native | 6 |
| Native Hawaiian or Other Pacific Islander | 6 |
| Other / Unknown | 256 |
| **5-Alpha-Reductase Inhibitor (5-ARI) Usage** |  |
| 5-ARIs currently prescribed during MRI scan | 84 |
| 5-ARIs previously taken (>6 months before MRI/Date Unknown) | 143 |
| **Scanner/Protocol Group Cohorts** |  |
| Cambridge | 5 |
| MGH | 63 |
| UCSD CTIPM<sub>Discovery1</sub>/UCSDH<sub>Discovery</sub> | 844 |
| UCSD CTIPM<sub>Discovery2</sub> | 167 |
| UCSDH<sub>Premier</sub> | 143 |
| UCSF/UCSD CTIPM<sub>Premier</sub> | 220 |
| UTSA<sub>Skyra</sub> | 55 |
| URMC | 251 |
| UTSA<sub>Trio</sub> | 88 |
| Other | 54 |

Statistically significant effects (*p < 0.05*) on RSIrs were observed using both Method 1 and Method 2 for age and for three acquisition groups (UCSD CTIPM_Discovery2_, URMC, and UTSA_Trio_) (Table 2). Three more acquisition groups had statistically significant effects using Method 1 but not Method 2 (UCSF/UCSD CTIPM_Premier_, UCSDH_Premier_, and UTSA_Skyra_). Prostate volume had a statistically significant effect on RSIrs_max_ using Method 2 but not Method 1. As expected, GG ≥2 had a significant statistical effect on RSIrs_max_. We repeated the Method 2 analysis using the 99^th^ percentile RSIrs due to presumed decreased variability/noise compared to the 100^th^ percentile (RSIrs_max_) and obtained similar results (Supplementary Table 2). Statistically significant effects of patient factors age and prostate volume were small, relative to the differences in RSIrs_max_ between patients with and without csPCa.

**Table 2.**
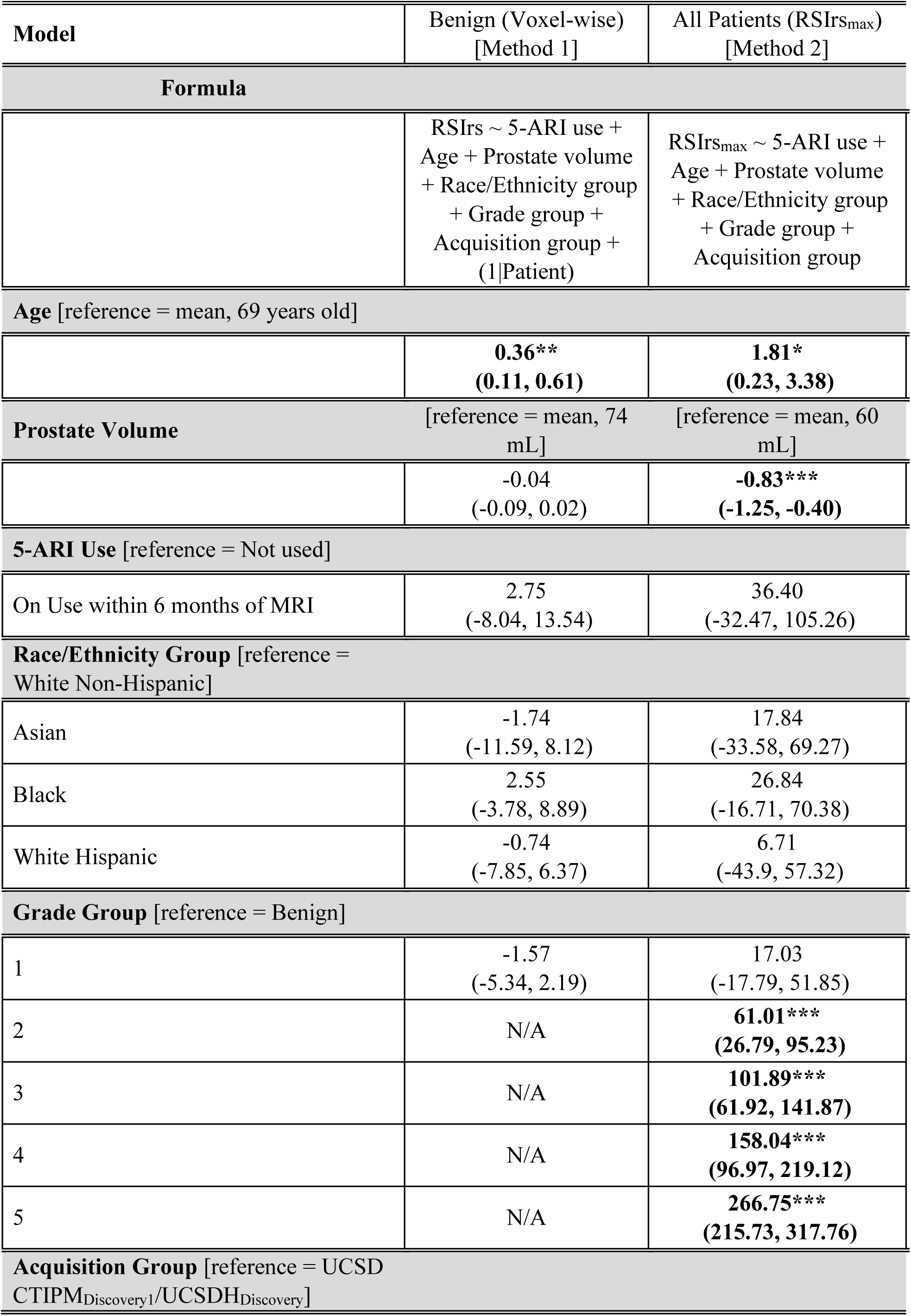
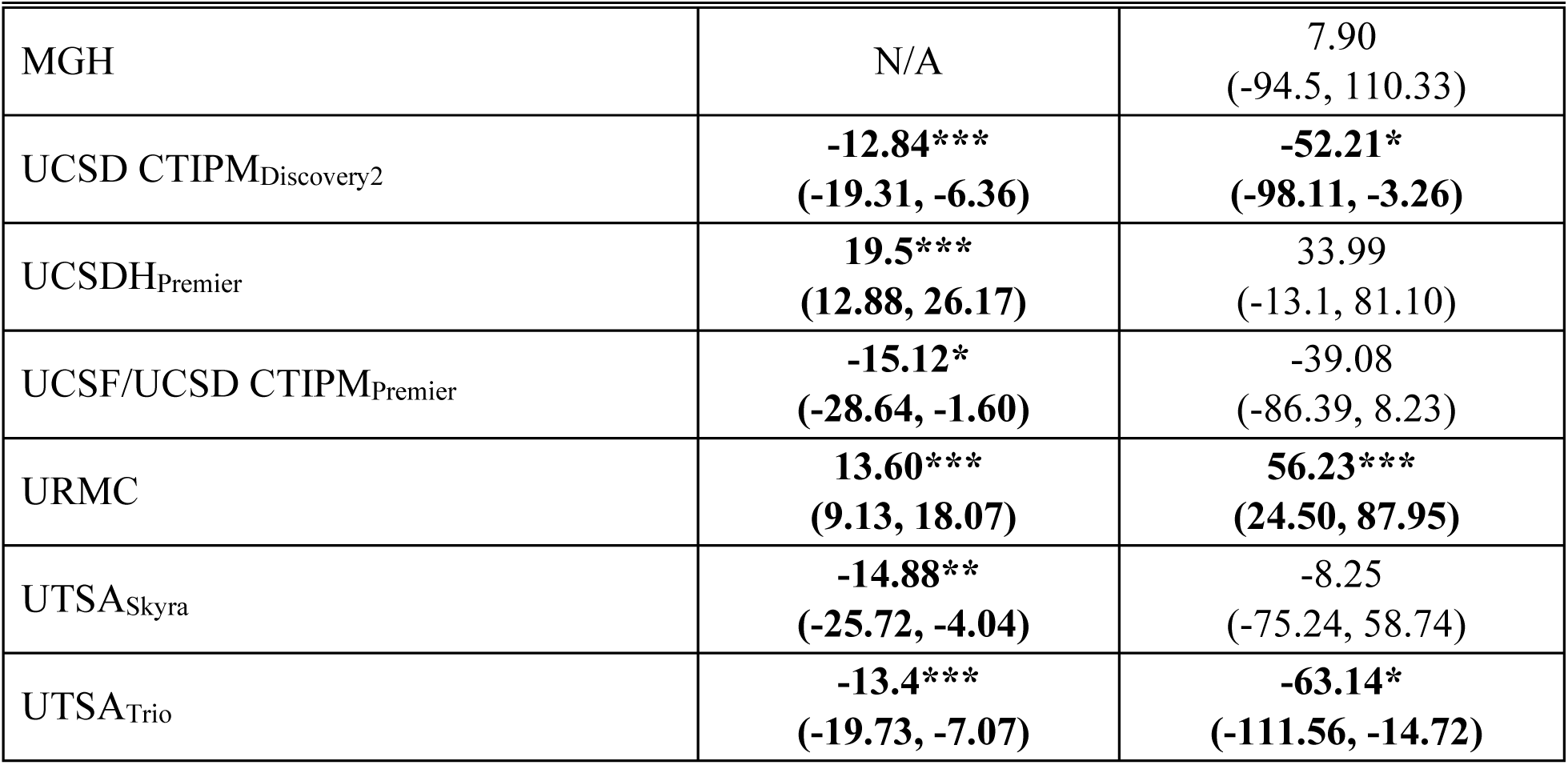
All predictors and their estimated effects on the RSIrs biomarker identified using linear mixed effects modeling [Method 1] and multiple linear regression modeling [Method 2]. These predictors included 5-ARI (current 5-ARI usage or usage <6 months before MRI), age, prostate volume, race/ethnicity, grade group, and acquisition group. Linear mixed effects modeling considered multiple voxels within the same patient. Coefficient estimates (95% confidence interval) are reported for each significant effect, which provide insight into the impact of these variables on RSIrs. *N/A* signifies there were no representative patients of that category included in the analysis. Significant predictors: * (*p < 0.05), ** (p < 0.01), *** (p < 0.001)*

Considered alone, adjustment appears to increase overlap between RSIrs distributions of effect groups to the reference, but this did not significantly increase similarity (Figure 3). Adjustment for patient and acquisition factors did not improve detection of csPCa with RSIrs (*p ≥ 0.05*) (Table 3). Pre-adjustment AUC was 0.77 [95% CI: 0.75-0.79]; post-Adjustment AUC was 0.77 [95% CI: 0.76-0.79] and 0.74 [95% CI: 0.72-0.76] using Method 1 and 2, respectively (Figure 4). A secondary analysis using patient subgroups selected specifically for the statistically significant factors using only Method 1 or only Method 2 similarly revealed no improvement of AUC when adjusting for factors with either approach (Supplementary Table 3).

**Figure 3.**
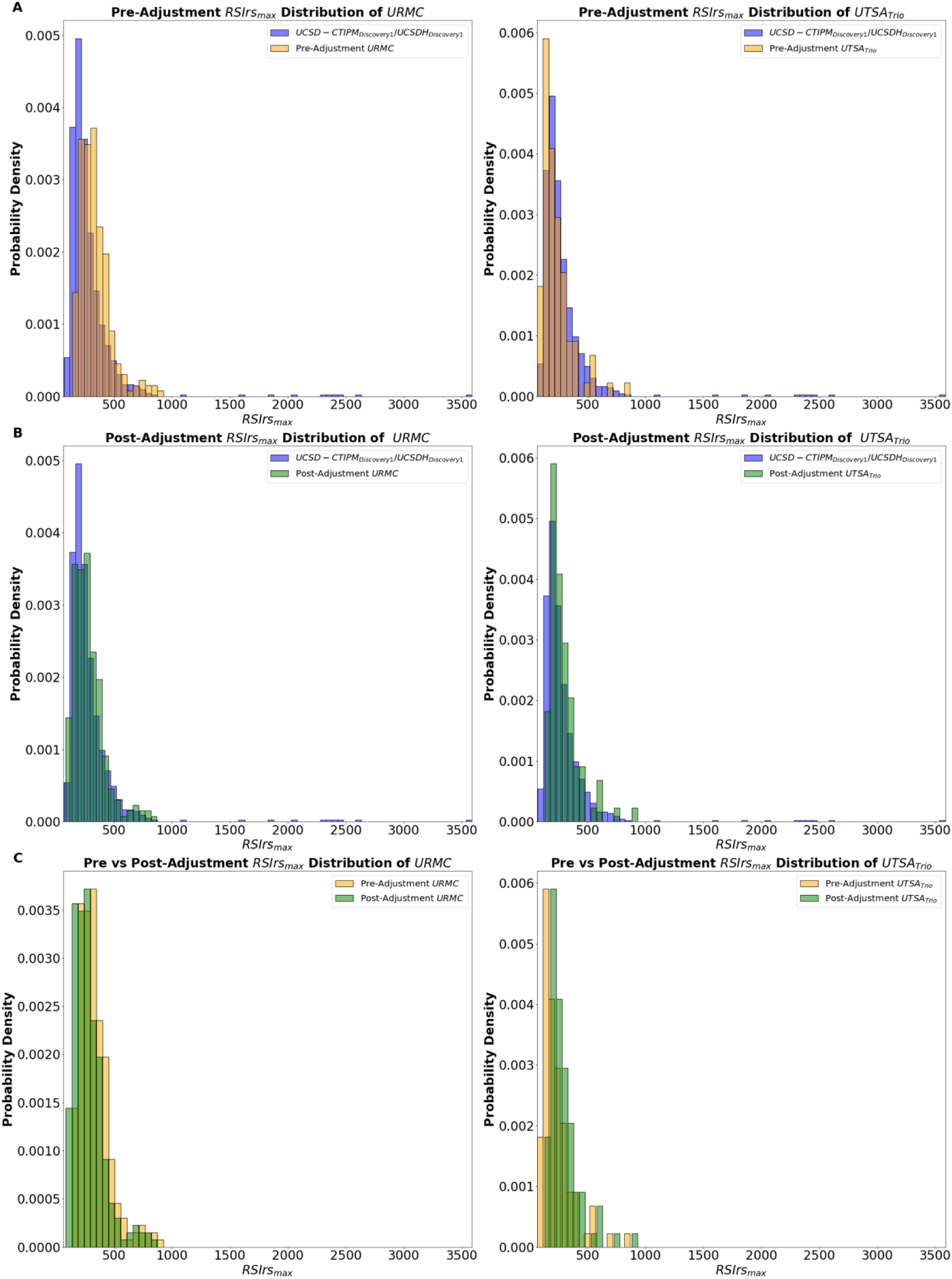
Histograms comparing the pre-adjustment and post-adjustment RSIrs distributions for two institutions with significant acquisition effects, URMC and UTSA_Trio_ (respectively in each, the orange and green histograms), using adjustments from Method 2 and the reference group UCSD CTIPM_Discovery1_/UCSDH_Discovery_ (blue histogram) are shown in plots (A) and (B). Plot (C) shows the pre-and post-adjustment RSIrs within each acquisition group. The distributions are significantly different (*p < 0.05)* before and after adjustment with Mann-Whitney U testing.

**Figure 4.**
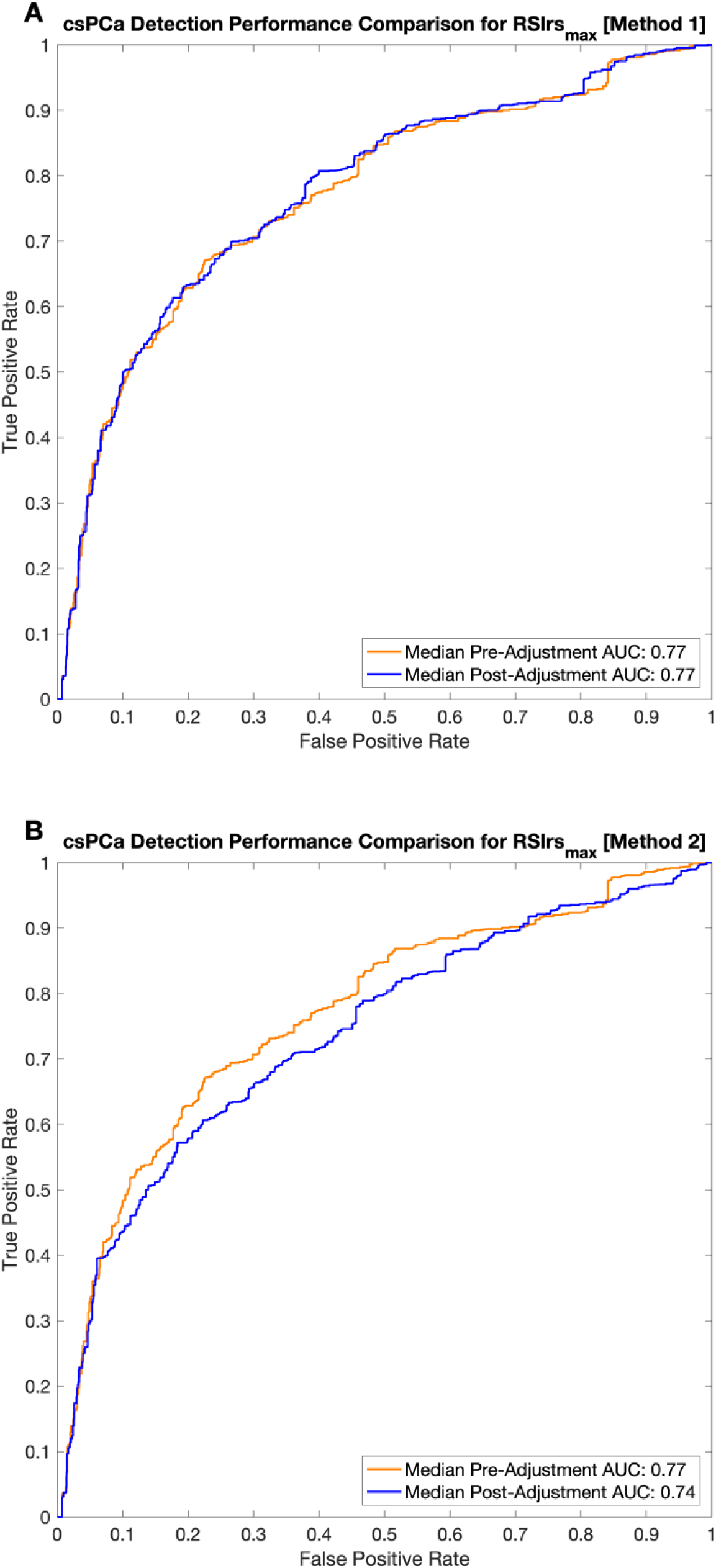
ROC curves illustrate the detection performance of RSIrs_max_ for clinically significant prostate cancer (csPCa) pre-adjustment and post-adjustment using Method 1 (Plot A) and Method 2 (Plot B). Pre-adjustment RSIrs_max_ performance (orange line) is compared with post-adjustment performance (blue line) after pooling data from 10,000 bootstrap samples. The area under the curve (AUC) values are reported for each model, demonstrating the impact of acquisition and patient adjustments on the predictive accuracy of RSIrs_max_ for csPCa. Median pre-adjustment AUC was 0.77 [95% CI: 0.75-0.79]. For Method 1, median post-adjustment AUC was 0.77 [0.76-0.79]. For Method 2, median post-adjustment AUC was 0.74 [0.72-0.76]. Adjustment for patient and acquisition effects does not significantly affect the AUC (*p < 0.05*).

**Table 3.** Results from a 10,000-bootstrap analysis using a subgroup of patients with significant acquisition and patient effects on RSIrs from both estimation methods. Each patient was matched with one in the reference population, stratified by grade group. A bootstrap sample size of 1000 was used. Adjustments were made using a linear transformation based on significant effects identified by each model, allowing comparison of AUC values pre-and post-adjustment. Adjusting for patient and acquisition effects did not improve csPCa detection using RSIrs_max_ (*p ≥ 0.05)*, suggesting the statistically significant effects on RSIrs_max_ in this cohort may be too small to affect the clinical utility of the imaging biomarker.

| Model used for Adjustments | Median AUC (Pre-Adjustment) | Median AUC (Post-Adjustment) | Median AUC Difference |
| --- | --- | --- | --- |
| Voxel-wise model in patients without csPCa [Method 1] | 0.77<br>[0.75, 0.79] | 0.77<br>[0.76, 0.79] | 0.005<br>[0.004, 0.006] |
| Patient-level model of RSIRS<sub>max</sub> using all patients [Method 2] |  | 0.74<br>[0.72, 0.76] | -0.030<br>[-0.036, -0.022] |

Estimation of significant factor effects appeared stable with around 20 patients typically adequate for stable estimation of effects of an acquisition group from a different scanner manufacturer and RSI protocol (URMC) to the reference population (Supplementary Figure 1).

## Discussion

Some patient and acquisition factors were associated with statistically significant systematic changes in RSI, on average, but these effects were small. Previous work has shown that RSIrs_max_ performs comparably to expert PI-RADS interpretation^14,16^. Here, we find that adjusting for potential patient and acquisition bias did not improve csPCa detection performance with RSIrs_max_. In other words, the present results suggest adjustment may be unnecessary, and RSIrs_max_ can be used effectively without major concern that age, self-reported race or ethnicity, prostate volume, or use of 5-ARIs will invalidate quantitative results. Likewise, our analysis of systematic “batch effects” from different scanner and acquisition protocol parameters suggests RSIrs is reliable across a range of centers, scanners, vendors, and acquisition protocols. These findings are consistent with the strong performance of RSIrs for csPCa detection in a recent study pooling heterogeneous data^16^. Here, we also demonstrate that the strongest factor impacting RSIrs_max_ in multivariable models is PCa GG, which is precisely the primary goal in developing a quantitative biomarker for csPCa.

Systematic biases can arise in laboratory values and biomarkers because of variation in the specific lab array, and any number of patient factors^37,38^. Measuring these potential biases is key to ensuring accurate and equitable utility of any biomarker. The small effects observed in our study are reassuring, as are the results of the sample-size analysis, which suggest 20 patients can be adequate to estimate systematic deviation of a new population or new factor from a reference population. Overall, this study demonstrates the robustness of RSIrs as a reproducible biomarker.

Prostate mpMRI has wide variation in performance in terms of positive predictive value (PPV)^39^. Quantitative biomarker development can improve reliability and reproducibility and is a stated priority of the NCI, NIBIB, RSNA, etc^40–43^. ADC is the quantitative marker currently used in mpMRI interpretation, but itperforms poorly as an objective biomarker in the absence of expert-defined suspicious lesions^14,16,40^. RSI and several other proposed advanced diffusion MRI models attempt to better explain the biophysical complexity of prostate tissue microstructure than ADC^17,44–58^. Each has shown improvements over conventional MRI, with a recent clinical trial finding a derived quantitative biomarker from Vascular, Extracellular, and Restricted Diffusion for Cytometry in Tumor (VERDICT) MRI, the fractional intracellular volume (FIC), as a superior classifier of csPCa^58^. A multi-center trial is also ongoing to evaluate the impact of RSIrs on accuracy of biopsy decisions by expert and non-expert radiologists (NCT06579417)^59^. To our knowledge, the present study is the largest investigation of reliability of a quantitative imaging biomarker for csPCa across commonly encountered patient factors and acquisition variability.

Limitations of this study included reliance on interpretation with PI-RADS and on biopsy results, both of which are subject to inter-reader variability, though our approach mirrors the reality of clinical practice. It is also possible that non-linear modeling of patient or acquisition factors could be more effective than linear models for assessing systematic effects, and this is left to future work. On the other hand, performance for csPCa detection with RSIrs is already sufficient for clinical utility with the implementation of voxel-wise RSIrs overlays providing benefit for contouring lesions without expert radiologists^14,16^. We note that some of the patient factors are found in only a minority of patients analyzed here such as 5-ARI usage or Black race. Larger studies may prove informative, but it is reassuring that the sample-size analysis demonstrates a plateau for the factors studied, including Black race. Finally, the systematic effects measured here represent correlations and may not imply causality; patients from a given imaging center may also simply differ from other populations in ways not measurable in the present work. For acquisition factors, it is possible to scan the same patient with both approaches—work we have done and are continuing to do^30^. For most patient factors (age, race, prostate volume, 5-ARI use), though, re-scanning without the factor is not a feasible strategy.

## Conclusion

RSIrs appears robust to many patient and acquisition factors, contributing to its potential as a quantitative imaging biomarker for csPCa.

## Data Availability

All data produced in the present study are available upon reasonable request to the authors

