## Supplementary Figures for "Robustness of a Restriction Spectrum Imaging (RSI) quantitative MRI biomarker for prostate cancer: assessing for systematic bias due to age, race, ethnicity, prostate volume, medication use, or imaging acquisition parameters"

| Institution | Scanner Models | Number of Scanners |
|---|---|---|
| UCSD CTIPM | GE Healthcare Discovery MR750,<br>GE Healthcare Signa Premier | 4 |
| UCSD Health | GE Healthcare Discovery MR750,<br>GE Healthcare Signa Premier | 4 |
| URMC | SIEMENS Magnetom Skyra | 2 |
| MGH | GE Healthcare Signa Premier | 1 |
| UCSF | GE Healthcare Signa Premier | 2 |
| Cambridge | GE Healthcare Discovery MR750 | 1 |
| UTHSCSA | SIEMENS Magnetom Skyra,<br>SIEMENS Magnetom Trio | 3 |

| Cambridge | DWI | T2-weighted |
|---|---|---|
| Pulse sequence | Diffusion-weighted EPI | Fast Spin Echo (FSE) |
| TR (ms) | 4500 | 3130 |
| TE (ms) | 59 | 98 |
| Voxel Size (mm) | 2.29 x 2.29 x 4 | 0.4 x 0.7 x 3 |

| | | |
|---|---|---|
| *b*-values (s/mm^2) [number of samples] | 0[1], 500[6], 1000[6], 2000[12] | N/A |
| Field Strength (T) | 3 | 3 |
| Scanner Manufacturer/Model | GE Healthcare/Discovery MR750 | GE Healthcare/Discovery MR750 |
| **MGH** | **DWI** | **T2-weighted** |
| Pulse sequence | Diffusion-weighted EPI | Fast Spin Echo (FSE) |
| TR (ms) | 4500 | 3937 |
| TE (ms) | 59 | 169 |
| Voxel Size | 2.5 x 2.5 x 6 | 0.44 x 0.71 x 3 |
| *b*-values (s/$mm^2$) [number of samples] | 0[1],500[8],1000[8],2000[16] | N/A |
| Field Strength (T) | 3 | 3 |
| Scanner Manufacturer/Model | GE Healthcare/Signa Premier | GE Healthcare/Signa Premier |
| **UCSD CTIPM_Discovery1/UCSDH_Discovery1** | **DWI** | **T2-weighted** |
| Pulse sequence | Diffusion-weighted EPI | Fast Spin Echo (FSE) |
| TR (ms) | 4500 | 7000 |
| TE (ms) | 69 | 100 |
| Voxel Size (mm) | 2.5 x 2.5 x 6 | 0.75 x 0.75 x 3 |
| *b*-values (s/mm^2) [number of samples] | 0[1], 500[6], 1000[6], 2000[12] | N/A |
| Field Strength (T) | 3 | 3 |
| Scanner Manufacturer/Model | GE Healthcare/Discovery MR750 | GE Healthcare/Discovery MR750 |
| **UCSD CTIPM_Discovery2** | **DWI** | **T2-weighted** |
| Pulse sequence | Diffusion-weighted EPI | Fast Spin Echo (FSE) |
| TR (ms) | 4500 | 6230 |
| TE (ms) | 76 | 98 |
| Voxel Size (mm) | 2.5 x 2.08 x 3 | 0.75 x 0.75 x 3 |
| *b*-values (s/$mm^2$) [number of samples] | 0 [1], 50[6], 800[6], 1500[12], 3000 [18] | N/A |
| Field Strength (T) | 3 | 3 |
| Scanner Manufacturer/Model | GE Healthcare/Discovery MR750 | GE Healthcare/Discovery MR750 |
| **UCSDH_Premier** | **DWI** | **T2-weighted** |
| Pulse sequence | Diffusion-weighted EPI | Fast Spin Echo (FSE) |
| TR (ms) | 4500 | 3423 |
| TE (ms) | 59 | 170 |
| Voxel Size (mm) | 1.9656 x 2 x 4 | 0.45 x 0.7 x 3 |

| | | |
|---|---|---|
| *b*-values ($s/mm^2$) [number of samples] | 0[1], 500[6], 1000[6], 2000[12] | N/A |
| Field Strength (T) | 3 | 3 |
| Scanner Manufacturer/Model | GE Healthcare/Signa Premier | GE Healthcare/Signa Premier |
| **UCSF/UCSD CTIPM_Premier** | **DWI** | **T2-weighted** |
| Pulse sequence | Diffusion-weighted EPI | Fast Spin Echo (FSE) |
| TR (ms) | 4500 | 2964 |
| TE (ms) | 75 | 150 |
| Voxel Size (mm) | 2.5 x 2.5 x 3 | 0.43 x 0.43 x 3 |
| *b*-values ($s/mm^2$) [number of samples] | 0[5], 100[6], 800[12], 1400[12], 2500[18] | N/A |
| Field Strength (T) | 3 | 3 |
| Scanner Manufacturer/Model | GE Healthcare/Signa Premier | GE Healthcare/Signa Premier |
| **URMC** | **DWI** | **T2-weighted** |
| Pulse sequence | Diffusion-weighted EPI | Fast Spin Echo (FSE) |
| TR (ms) | 3800 | 4800 |
| TE (ms) | 85 | 104 |
| Voxel Size (mm) | 1 x 1 x 4 | 0.47 x 0.49 x 3 |
| *b*-values ($s/mm^2$) [number of samples] | 0[1], 500[6], 1000[6], 2000[6] | N/A |
| Field Strength (T) | 3 | 3 |
| Scanner Manufacturer/Model | SIEMENS Healthcare/Magnetom Skyra | SIEMENS Healthcare/Magnetom Skyra |
| **UTSA_Skyra** | **DWI** | **T2-weighted** |
| Pulse sequence | Diffusion-weighted EPI | Fast Spin Echo (FSE) |
| TR (ms) | 6300 | 4710 |
| TE (ms) | 105 | 100 |
| Voxel Size (mm) | 1 x 1 x 4 | 0.75 x 0.56 x 3 |
| *b*-values ($s/mm^2$) [number of samples] | 0[1], 500[6], 1000[18], 2000[42] | N/A |
| Field Strength (T) | 3 | 3 |
| Scanner Manufacturer/Model | SIEMENS Healthcare/Magnetom Skyra | SIEMENS Healthcare/Magnetom Skyra |
| **UTSA_Trio** | **DWI** | **T2-weighted** |
| Pulse sequence | Diffusion-weighted EPI | Fast Spin Echo (FSE) |
| TR (ms) | 98 | 1800 |
| TE (ms) | 5500 | 203 |
| Voxel Size (mm) | 2.3438 x 2.3438 x 3 | 0.68 x 0.68 x 1.5 |

| | | |
|---|---|---|
| *b*-values (s/mm$^2$) [number of samples] | 0[1], 500[30], 1000[30], 2000[30] | N/A |
| Field Strength (T) | 3 | 3 |
| Scanner Manufacturer/Model | SIEMENS Healthcare/Magnetom Trio | SIEMENS Healthcare/Magnetom Trio |

Supplementary Table 1. Detailed Breakdown of MRI parameters for all acquisition groups. *N/A* signifies the parameter was not relevant for the acquisition. Parameters may differ between patient MRI scans. Abbreviations: TR (Repetition Time), TE (Echo Time), FSE (Fast Spin Echo), EPI (Echo Planar Imaging), Cambridge ( University of Cambridge), MGH (Harvard University's Massachusetts General Hospital), UCSD[H] (University of California San Diego [Health]), CTIPM (Center for Translational Imaging and Precision Medicine), UCSF (University of California San Francisco), URMC (University of Rochester Medical Center), UTHSCSA (University of Texas Health Sciences Center San Antonio).

| Model | All Patients ($RSIrs_{99}$) |
|---|---|
| **Formula** | |
| | $RSIrs_{99}$ ~ 5-ARI use + Age + Prostate volume + Race/Ethnicity group + Grade group + Acquisition group |
| **Age** [reference = mean, 69 years old] | |
| | **1.07*<br>[0.04, 2.11]** |
| **Prostate Volume** [reference = mean, 60 mL] | |
| | **-0.80***<br>[-1.08, -0.52]** |
| **5-ARI Use** [reference = Not used] | |
| On Use within 6 months of MRI | 29.69<br>[-15.78, 75.16] |
| **Race/Ethnicity Group** [reference = White Non-Hispanic] | |
| Asian | 14.25<br>[-19.70, 48.20] |
| Black | 6.51<br>[-22.25, 35.26] |
| White Hispanic | -13.34<br>[-46.76, 20.07] |
| **Grade Group** [reference = Benign] | |
| 1 | 8.43<br>[-16.11, 33.01] |
| 2 | **31.77**<br>[9.18, 54.36]** |
| 3 | **56.51***<br>[30.12, 82.91]** |
| 4 | **112.78***<br>[72.451, 153.10]** |
| 5 | **182.8***<br>[149.12, 216.48]** |
| **Acquisition Group** [reference = UCSD CTIPM_Discovery1] | |
| MGH | 43.51<br>[-24.12, 111.14] |

| | |
|---|---|
| UCSD CTIPM$_{Discovery2}$ | **-68.54***<br>[-98.85, -38.23]** |
| UCSDH$_{Premier}$ | 29.12<br>[-1.99, 60.22] |
| UCSF/UCSD CTIPM$_{Premier}$ | **-41.67**<br>[-72.9, -10.43]** |
| URMC | **33.33**<br>[12.39, 54.28]** |
| UTSA$_{Skyra}$ | -39.66<br>[-83.89, 4.57] |
| UTSA$_{Trio}$ | **-45.05**<br>[-77.01, -13.07]** |

Supplementary Table 2. All predictors and their estimated effects on the 99$^{th}$ percentile RSIrs identified using multiple linear regression modeling [Method 2]. These predictors included 5-ARI (current 5-ARI usage or usage <6 months before MRI), age, prostate volume, race/ethnicity, grade group, and acquisition group. Coefficient estimates [95% confidence interval] are reported for each significant effect, which provide insight into the impact of these variables on RSIrs. *N/A* signifies there were no representative patients of that category included in the analysis. Significant predictors: * (*p < 0.05*), ** *(p < 0.01)*, *** *(p < 0.001)*

| **Model used for Adjustments** | **Median AUC (Pre-Adjustment)** | **Median AUC (Post-Adjustment)** | **Median AUC Difference** |
|---|---|---|---|
| Voxel-wise model in patients without csPCa [Method 1] | 0.76 [0.74, 0.78] | 0.77 [0.75, 0.79] | 0.007 [0.006, 0.009] |
| Patient-level model of $RSIrs_{max}$ using all patients [Method 2] | 0.74 [0.72, 0.76] | 0.73 [0.70, 0.75] | -0.01 [-0.02, -0.005] |

Supplementary Table 3. Results from two 10,000-bootstrap analyses using subgroups of patients with significant acquisition and patient effects on RSIrs estimated with two methods, respectively. Each patient was matched with one in the reference population, stratified by grade group. A bootstrap sample size of 1000 was used. Adjustments were made using a linear transformation based on significant acquisition and patient effects identified by two different estimation methods, allowing comparison of AUC values pre- and post-adjustment. This reflects the impact of both acquisition and patient effects.

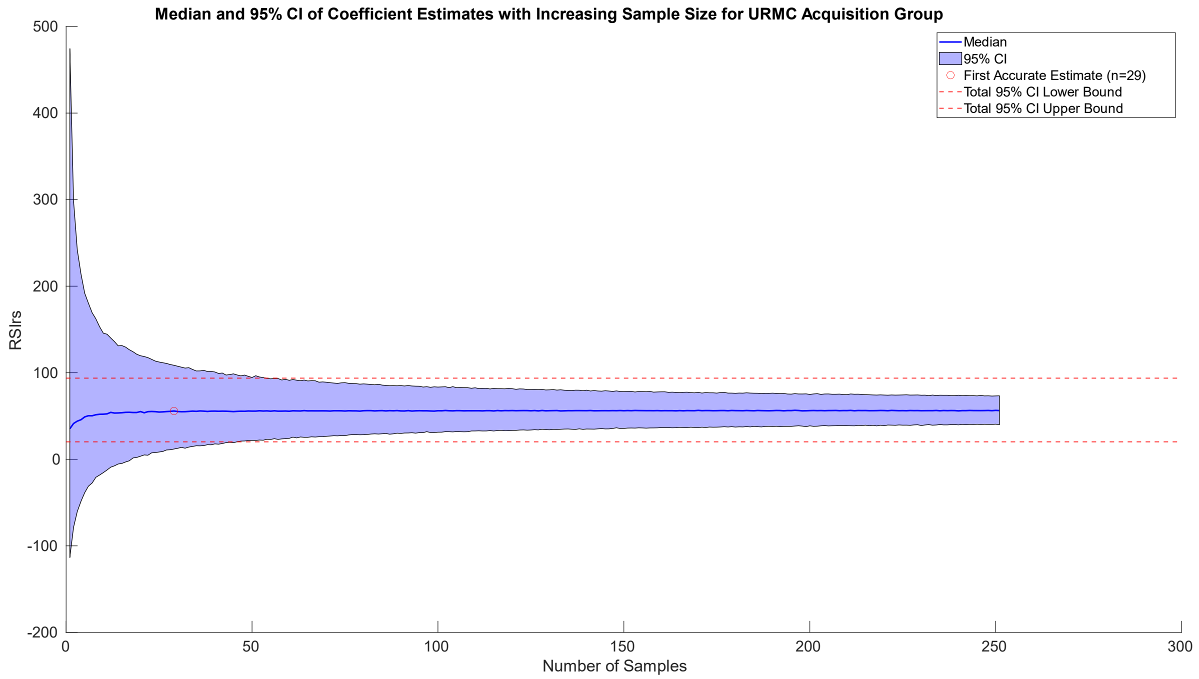


Supplementary Figure 1. Sample size estimation analysis for an acquisition group from University of Rochester Medical Center (URMC). In this analysis, 10,000 bootstrap samples ranging from n = 1:251 were taken. The line on the graph represents the median effect estimate for each set of 10,000 bootstrap samples (RSIrs), while the blue shading indicates the 95% Confidence Interval for each set of 10,000 bootstrap samples. The red dotted lines represent the mean 95% Confidence Interval of all sets of bootstrap samples. The red circle denotes the location of the minimum estimated sample size required to accurately reproduce the acquisition effects at this institution, defined as where the minimum bootstrap median coefficient estimate that is within 1% of the value observed for the total population (n=251). Accurate estimation of acquisition effects requires only 29 patients.
